# Evaluating the clinical effectiveness and implementation outcomes of the nurse-led stroke transition care model in Tanzania: a study protocol for effectiveness-implementation design

**DOI:** 10.1101/2025.06.17.25329815

**Authors:** Nyagwaswa Athanas Michael, Lilian Teddy Mselle, Costansia Anselim Bureta, Yingjuan Cao

## Abstract

**Background:** Stroke remains a leading cause of death and disability worldwide, imposing a significant burden on individuals, and healthcare systems. Globally, low- and middle-income countries (LMICs) bear over 80% of stroke-related deaths that occur soon after hospital discharge. This highlight an urgent need for a well-coordinated, transition care model that ensures continuity of care, supports patients and caregivers at home, and bridges the critical gap between hospital discharge and home care. The current study aims to evaluate the clinical effectiveness and implementation outcomes of a nurse-led stroke transition care model within the Tanzania’s healthcare system.

**Materials and methods:** This study adopts an effectiveness-implementation design hybrid type 1 with mixed-methods approach. About 77 pairs stroke survivors and caregivers, as well as 77 pairs of clinical nurses and physicians will be recruited in the pre-implementation phase and implementation phase from September 2024 to March 2025, and June to December, 2025 respectively at Muhimbili National Hospital-Mloganzila, a national stroke center in Tanzania. Descriptive statistics will be conducted using Independent Samples T-test, One-way ANOVA, Chi-Square Test (X^2^), Fisher’s Exact Test, Linear Mixed Models, Kaplan-Meier and Cox regression using STATA software. Furthermore, audio-recorded in-depth interviews will be conducted from November to December, 2025 among 10-20 triads of stroke survivors, caregivers and healthcare providers, to evaluate the implementation outcomes of the model. Transcribed data will be entered in Dedoose software for analysis using a thematic analysis with an inductive approach. The study has obtained approval from Muhimbili University of Health and Allied Sciences Institution Review Board (MUHAS-REC-04-2024-2139).

**Conclusion:** Given the rising burden of stroke and the gaps in continuity of care after hospital discharge, implementing a structured, nurse-driven approach could enhance patient outcomes, support caregivers, and reduce the risk of complications or readmissions.

## 1.0 Introduction

Stroke is a second leading cause of death and disability worldwide, with ischemic stroke accounting for more than half of all stroke types (1). The global prevalence, incidence and mortality rates of stroke are estimated to be 101 million, 12.2 million and 6.6 million respectively in 2019. On the same year, the disability adjusted life-years (DALYs) from stroke was estimated to be 143 million. This represents a substantial rise in stroke burden for 85% prevalence rate, 70% incidence rate, 43% death rate, and 32% DALYs in the past two decades (2). The observed stroke vital statistics are two times higher in Sub-Sahara African countries than those reported in high-income countries (2).

Therefore, there is a growing need to understand the burden of stroke and its associated treatment outcomes in Africa, a continent facing unique healthcare challenges. Stroke prevalence and incidence in Africa exhibits considerable variability, with a higher burden in Sub-Saharan Africa (SSA). Studies have reported the pooled crude incidence rate and one-month case-fatality rates of 106.49 per 100,000 and 24.5% respectively in SSA (3). While in Northern Africa the age-standardized point prevalence, death and DALY rates of stroke are estimated to be 1537.5, 87.7 and 1826.2 per 100,000 respectively (4).

Limited access to stroke care services, late or underdiagnoses, and inadequate stroke-capable facilities remain the significant predictors of mortality due to stroke in Africa (5). Addressing the stroke burden in the continent requires a multifaceted approach that involve preventive strategies targeting risk factors, improving access to stroke care, establishing stroke units, and increasing rehabilitation services (6). Interestingly, number of efforts are underway in several countries to raise awareness about stroke risk factors, promote healthy lifestyles, and improving access to acute stroke care (7). However, efforts to address transition care (TC) have not been widely realized in the current initiatives.

Transitioning from hospital to home is a critical phase for patients with chronic illnesses such as stroke. Stroke, being a sudden and often life-altering condition, requires a comprehensive TC approach to facilitate recovery, community reintegration, and long-term disease self-management (8). Owing the growing stroke burden in low-and-middle income countries (LMICs), it is crucial for the healthcare systems to establish a well-functioning TC pathway to enhance quality of stroke care. Lack of a seamless transition may leave stroke survivors susceptible to readmissions and suboptimal health outcomes (9).

The current TC for patients with stroke in SSA countries such as Tanzania is fraught with multifaceted challenges. Stroke survivors are discharged directly from hospital to home leading to a shift of locus of care from healthcare providers to untrained caregivers. Sometimes, hospital to home discharge is a surprise to stroke survivors and their caregivers because they are seldom involved in discharge planning. Furthermore, the transition process lacks a cohesive coordination among healthcare providers resulting in disjointed care, where essential post-stroke care and rehabilitation services are missed (10).

To improve TC, several strategies need to be piloted. First, establishing standardized protocols for information exchange between stroke survivors, caregivers and healthcare providers to ensure timely and complete transfer of patient data. Second, involving stroke survivors and their caregivers in discharge planning, offering clear and understandable instructions, and providing educational resources to empower patients to take an active role in managing their health. Third, designing and implementing locally validated evidence-based TC models to improve hospital-to-home transition. Since plenty of evidence is available in-favor of nurse-led stroke TC (11), adoption of these evidence-based models is worth to explore within the Tanzania’s healthcare system.

The current study aims to evaluate the clinical effectiveness and implementation outcomes of a locally validated nurse-led stroke TC model among stroke survivors, caregivers and healthcare providers in Tanzania. Ideally, the nurse-led stroke TC model is informed by the philosophy of the end-user also known as user-centered design (12). The model intends to; (a) prepare stroke survivors to return home through involving them in discharge planning, and (b) foster a sense of patient self-efficacy through active participation in disease self-management.

## 2.0 Materials and methods

### 2.1 Study design

This study uses a multi-stage mixed-methods approach with an effectiveness-implementation design hybrid type 1 that focuses predominantly on clinical effectiveness, and minimal implementation outcomes of the nurse-led stroke TC model (13).

### 2.2 Setting

The study will be conducted in Dar es Salaam: the most urbanized and commercial city in Tanzania, with a population of about 5.4 million people (14). Specifically, the study will be conducted at stroke units and neurology wards of Muhimbili National Hospital (MNH)-Mloganzila located at Ubungo district. The hospital has a bed capacity of 600 with an internal medicine department that has 108 beds (15). MNH-Mloganzila is a national stroke care center in Tanzania. About 50 stroke survivors are discharged from the stroke units per month after receiving specialty care from neurologists, trained stroke nurses, clinical pharmacists, clinical nutritionists and rehabilitation therapists (15).

### 2.3 Inclusion and exclusion criteria

The study will include clinical nurses and physicians with six months of working experience in stroke care, and holding a minimum of diploma in their respective professions. Patients will be included given that they are 18 years old and above, admitted in the stroke units, primary diagnosis of stroke confirmed by brain CT/MRI, undergo usual discharge process, lives with a caregiver, has a mobile phone, can read and write, able to communicate, National Institutes of Health Stroke Scale (NIHSS) <5, Modified Barthel Index (MBI) > 10, Modified Rankin Scale (mRS) <4, Montreal Cognitive Assessment Test (MoCA) >20, expected to stay in the ward for 3-5 days, and expected to survive for 3 months (16). While, caregivers will be recruited given that they have a mobile phone, can read and write, able to communicate, and live with the patient after stroke. Healthcare providers who will be on leave during the study period will be excluded from the study. While, stroke survivors with previous stroke who are not admitted in stroke units, discharged against medical advice, have end-stage organ failure, without a caregiver, who can’t read/write, and without mobile phone will be excluded. Whereas, caregivers without mobile phone that is accessible will be excluded (16).

### 2.4 Development and validation of the Nurse-led Stroke Transition Care Model

The research team developed the nurse-led stroke TC model by conducting a systematic review and meta-analysis of randomized controlled trials, and rated the evidence by using GRADE recommendations (17). Then, in-depth interviews (IDIs) were conducted between June and September 2024 (Supplementary File 1) with a purposively sample of stroke survivors (n=15), caregivers (n=15) and healthcare providers (n=15). The qualitative phase aimed to explore barriers, facilitators and strategies to improve the current hospital-to-home stroke TC practices at the local setting. Findings from the systematic review and IDIs were combined to form an initial draft of the nurse-led stroke TC model containing TC interventions and nursing activities. Later, the nurse-led stroke TC model interventions and nursing activities were validated by using co-designing approach through three iterative Delphi rounds with academic experts, clinical experts, patients and informal caregivers between January and March, 2025 (18). Finally, the research team developed and validated the educational materials from literature review of post-stroke guidelines and expert opinions on discharge and post-discharge care of stroke survivors and their caregivers.

### 2.5 Feasibility of the Nurse-led Stroke Transition Care Model

Before implementation, we conducted a one-week in-housel training with healthcare providers on the locally validated nurse-led TC model. Healthcare providers were assessed their competences in providing TC through observer rating by the principal investigator. After one week of in-house training, a 4-week pilot of 5-pairs of stroke survivors and their caregivers was conducted from April to May, 2025. End-point outcome data were collected at 1 month. Assessment of perceived barriers and facilitators for implementing the nurse-led stroke TC was also conducted using the Consolidated Framework for Implementation Research (19). Likewise, the researchers assessed the acceptability, appropriateness and feasibility of the nurse-led stroke TC model by using the 20-items Implementation Outcome measures developed by Weiner and colleagues (20). Two clinical nurses with a minimum of bachelor degree were selected to be TC nurse champions who will implement the program in their respective units. Other two nurses with the same qualifications will be recruited from the academic institution.

### 2.6 Implementation of the Nurse-led Stroke Transition Care Model

The implementation of the nurse-led stroke TC model (Supplementary File 2) is informed by the Health Belief Model (HBM) and Social Cognitive Theory (SCT) (21). The model has ten interventions, each with two goals (Table 1). Two nurse champions will conduct the face-to-face sessions while other two nurse researchers will conduct the telephone call sessions (Table 2). Stroke survivors and their caregivers will be recruited to participate in the study upon admission in the stroke units from 1^st^ July to 30^th^ December, 2025. During hospitalization, the two nurse champions will conduct two individual sessions (at admission; session 1 and at discharge; session 5), and three group sessions (session 2, 3 and 4) with 2-4 dyads of patients and caregivers at the hospital conference rooms. After hospitalization, two nurse researchers will conduct seven follow-up sessions via phone calls at day 3, week 1, week 3, week 5, week 7, week 9, and week 11. The face-to-face sessions and telephone call sessions are expected to take 50-60 minutes and 20-30 minutes respectively.

**Table 1:** Transition Care Model Interventions and Goals.

| Transition care intervention elements | Goals |
| --- | --- |
| 1. Patient risk assessment | -To identify risks for hospitalization<br>-To identify risks for poor recovery |
| 2. Discharge planning | -To facilitate early discharge<br>-To formulate transition care goals |
| 3. Stroke specific education | -Identify stroke risk factors and warning signs<br>-Identify stroke complication<br>-To manage symptoms and seek care |
| 4. Disease self-management education | -To modify health risks such as eating, smoking, alcoholism and sleeping<br>-To develop culture of symptom self-assessment and management |
| 5. Caregivers education | -To improve caring willingness<br>-To enhance caregiver engagement in care |
| 6. Education on patient health record | -To enhance patient health literacy<br>-To facilitate care continuity and follow-up |
| 7. Teaching problem solving skills | -To improve social adaptability<br>-To develop coping skills |
| 8. Medication reconciliation | - To prevent medication error<br>-To enhance medication management |
| 9. Rehabilitation education | -To enhance adherence to scheduled physiotherapy sessions<br>-To equip caregivers with competence home-based exercises |
| 10. Education on follow-up care | -To complete treatment schedule<br>-Foster disease self-management |

**Table 2:** Nurse-Led Transition Care Model Implementation Algorithm.

| Transition care nursing activity | Hospitalization face-to-face sessions |  |  |  |  | Post-hospitalization phone call sessions |  |  |  |  |  |  |
| --- | --- | --- | --- | --- | --- | --- | --- | --- | --- | --- | --- | --- |
|  | 1 | 2 | 3 | 4 | 5 | 1 | 2 | 3 | 4 | 5 | 6 | 7 |
| -Conduct 8Ps of patient risk assessment<br>-Reconcile pre-admission and admission medications<br>-Conduct discharge home needs assessment<br>-Formulate individualized long- and short-term goals | √ |  |  |  |  |  |  |  |  |  |  |  |
| - Teach about stroke risk factors<br>- Teach about stroke warning signs<br>- Teach about stroke complication |  | √ |  |  |  |  |  |  |  |  |  |  |
| -Teach about symptom self-assessment<br>-Teach about managing anxiety and depression<br>-Teach about managing change of body image<br>-Teach about home-based physical exercises<br>-Teach about support systems |  |  | √ |  |  |  |  |  |  |  |  |  |
| -Teach how to solve and complex simple problems<br>-Educate on the importance of personal health records<br>-Teach about vital signs data |  |  |  | √ |  |  |  |  |  |  |  |  |
| -Assess completion of short-term goals<br>-Give customized discharge instructions<br>-Reinforce on behavioral and emotional management<br>-Teach on the need and importance of follow-up visit<br>-Teach patient about medication use and side effects |  |  |  |  | √ |  |  |  |  |  |  |  |
| -Discuss issues related to general health and home care<br>-Reinforce on stroke education<br>-Reinforce on self-management<br>-Reinforce caregivers' roles and responsibilities<br>-Reinforce on dealing and managing anxiety<br>-Reinforce on medication adherence<br>-Reinforce on maintaining personal health records<br>-Answer any questions related to follow-up visit |  |  |  |  |  | √ | √ | √ | √ | √ | √ | √ |

### 2.7 Intervention fidelity

The principal investigator will conduct face-to-face training and give printed materials and checklists to stroke TC program implementers to enhance intervention fidelity. Also the principal investigator will conduct participatory observations and peer ratings of implementers to assess adherence to the implementation algorithms. Implementers will be given feedback with regards to their adherence to implementation algorithm after finishing ward-rounds and clinical meetings. Poor adherence will imply a need for more education.

### 2.8 Sampling

The sample size is estimated by using statistical method with a G*power calculator to detect a mean difference between two groups for quality of life of stroke survivors as shown in a previous clinical trial of stroke transition care (22). The current study assumes a medium effect size of Cohen d = 0.5, 80% priori power, two tailed alpha = 0.05, and allocation ratio of 1:1. This yields a total sample size of 128. After accounting for 20% attrition rate, a minimum of 154 dyads of patients and caregivers will be required in pre-implementation phase also known as historical control group (n=77) and implementation phase also known as intervention group (n=77). Similarly, a minimum of 154 dyads of registered nurses and discharging physicians will be required to ascertain for patients’ discharge preparedness events in historical control group (n =77) and intervention group (n = 77) respectively.

### 2.9 Data collection

In the historical control group, data was collected between September 1, 2024 and March 30, 2025. However, in the intervention group, data will be collected between July and December, 2025. In each group, baseline data will be collected upon admission in the stroke units (T0). Follow-up will be done at 1 month (T1), 3 months (T2) and 6 months (T3) after discharge. Patients’ demographic characteristics will include age, sex, education level, insurance type, marital status, religion, and residence. Also, biodata such as blood pressure, blood sugar, lipid profile, body mass index, stroke type, comorbidity index, functional status, cognitive status, stroke severity, stroke onset, time metrics and medications will be collected. Caregivers’ demographic characteristics will include age, sex, education level, residence, phone number and relationship with patient. While, healthcare providers’ demographic characteristics will include age, sex, education level, profession, and years of work experience.

### 2.10 Evaluation of the Clinical Effectiveness of a Nurse-led Stroke TC Model

#### 2.10.1 Stroke survivors’ outcomes

The primary outcomes will be Quality of TC that will be measured by the Care Transitions Measure (CTM) Tool developed by Eric Coleman (23); Discharge preparedness that will be measured by the Short Forms of the Readiness for Hospital Discharge Scale (RHDS) developed by Weiss (24); Self-efficacy that will be measured by the Stroke Self-Efficacy Questionnaire (a=0.9) developed by Jones and colleagues (25); and Quality of life that will be measured by the Stroke Specific Quality of Life (SSQoL) with (a=0.85). The secondary outcomes will be resilience that is measured by the 10-items Connor–Davidson Resilience Scale (CD-RISC) validated by Laura (26) with a=0.85); depression and anxiety that is measured by the locally validated 10-items Kessler Psychological Distress Scale with a=0.85 (27); adherence to follow-up measured as number times a mandatory stroke visit is skipped; readmission measured as a number of times an unplanned readmission occurs; mortality measured as death that occurs after a patient is discharged from the hospital; and length of stay measured as number of days spent in the hospital from admission to discharge.

#### 2.10.2 Healthcare providers’ outcomes

The outcome for healthcare providers will be discharge readiness as measured by the Short Forms of the Readiness for Hospital Discharge Scale (RHDS) developed by Weiss (24).

#### 2.10.3 Caregivers’ outcomes

Caregiver self-efficacy that will be measured by the 10-items Family Caregiver Activation Tool (FCAT) developed by Coleman (28) with a=0.6; Resilience that will be measured by the 10-items Connor–Davidson Resilience Scale (CD-RISC) validated by Laura (26) with a=0.85); and Depression and anxiety that will be measured by the locally validated 10-items Kessler Psychological Distress Scale with a=0.85 (27).

#### 2.10.4 Data analysis

Quantitative data will be analyzed using STATA software. At baseline demographic characteristics, and group differences between the historical control group and intervention group will be analyzed by the Independent Samples T-test and one-way ANOVA. The Chi-Square Test of Associations (X^2^) and Fisher’s Exact Test will be conducted to compare association between categorical variables. Linear Mixed Models will be used to analyze repeated measures data, while modeling individual-averaged effects. To estimate, visualize and compare survival probabilities over time, Kaplan-Meier estimation and Log-rank test will be conducted. Also, Cox regression will be used to examine the relationships between predictor variables and the hazard rate or relative risk of experiencing an adverse event over time. A p < 0.05 and 95% Confidence Intervals (95% CI, excluding zero value) will be regarded as statistically significant indicators.

### 2.11 Evaluation of the Implementation Outcomes of the Nurse-led stroke TC Model

Descriptive qualitative explorations will be conducted to uncover the benefits, barriers and facilitators of implementing, adopting and sustaining the nurse-led stroke transition care model. Stroke survivors, caregivers and healthcare providers who participated during the implementation of the nurse-led stroke TC model will be will be purposeful selected (29) between November and December, 2025. Data collection will be done by using audio-recorded IDIs and Focus Group Discussions (FGDs). Initially 10-20 In-depth Interviews (IDIs) will be conducted, followed by 2-4 FGDs with 6-8 healthcare providers per group. Stroke survivors and caregivers will be recruited during their scheduled clinic visits at the out-patient department, while healthcare providers’ will be recruited from the neurological wards.

#### 2.11.1 Data analysis

Data analysis will be done using six-steps thematic analysis in Dedoose software. First, researchers will familiarize themselves with the data by reading transcripts multiple times. Second, one researcher competent in using the software will generate initial codes by identifying meaningful features in the data. Third, the researcher will search for themes by grouping related codes together. Fourth, themes will be reviewed to ensure they accurately represent the data. Fifth, the researchers will define and name each theme to clearly capture its essence. Finally, the research team will produce the report, where themes will be integrated into a coherent narrative that addresses the research questions. This qualitative phase will adhere to the established standards (credibility, confirmability, dependability, and transferability) for undertaking qualitative studies (30).

### 2.12 Quality control of the study

The study will use standardized and validated assessment tools that correspond to the study objectives and local context. Expert methodological opinions, suggestions, and recommendations have been solicited to improve the current study. Regular team meetings, discussions, and training before and after data collection will be hold. Any deviations from the study protocol will be timely communicated and rectified. Also, any missing data will be noted and measures to input data either directly by contacting the study participant or using multiple imputation method with chained equation will be considered.

### 2.13 Ethical considerations

The study adheres to the ethical principles of conducting studies involving human subjects as outlined in the Helsinki Declaration. The ethical approval to conduct this study has been obtained from Muhimbili University of Health and Allied Sciences Institution Review Board (MUHAS-REC-04-2024-2139). Permission to enter the study site has been granted by the Executive Director of MNH. All participants are required to give an informed written consent to participating in this study. The participants are informed about the purpose and benefits of participating in the study. Also. Participants are told that participation is voluntary and any person may wish to participate or drop out from the study at any time. Furthermore, participants’ information is kept under confidential and privacy grounds.

## 3.0 Discussion

Recently, nurse-led stroke TC models are increasingly recognized as effective strategies for improving various aspects of care, including discharge preparedness, and disease self-management (16). These TC models ensure that patients are adequately prepared for discharge. Nurses use their clinical expertise and patient-centered approach to assess patients’ needs, addressing concerns, and coordinating TC. During transition, nurses deliver comprehensive education and training to patients and their caregivers on various aspects of care such as self-care, medication management, symptom recognition, rehabilitation exercises, and lifestyle modifications (31).

The nurse-led TC models have been shown to positively impact the quality of life of stroke patients by addressing their physical, emotional, and social needs (32). Nurses play a crucial role in managing stroke-related symptoms and complications, such as pain, fatigue, depression, and anxiety. Through patient education, symptom monitoring, and timely interventions, nurses alleviate distressing symptoms and improve patients’ overall quality of life. Also, nurses provide psychosocial support and counseling to patients in order to cope with the psychological and emotional impacts of stroke (33).

Evidence show that nurse-led TC models not only have positive impacts on patients’ outcomes, but also have significant impacts on caregivers’ resilience, self-efficacy, and activation. TC nurses offer caregivers education and training on assisting patients with activities of daily living, managing medications, and recognizing signs of complications (32). Similarly, TC nurses enhance caregiver self-efficacy by providing opportunities for skill development, fostering confidence in caregiving abilities, and promoting a sense of mastery over caregiving tasks. Also, during transition, caregivers are engaged as active partners in the care process that promotes a sense of ownership over caregiving responsibilities (34).

Moreover, nurse-led TC models facilitate effective communication and collaboration among healthcare providers from different disciplines involved in the care of stroke survivors (8). These models emphasize care coordination among healthcare providers to ensure seamless transitions for stroke survivors. Nurses serve as central coordinators, liaising between different disciplines and facilitating the exchange of information and resources. By promoting regular team meetings, multidisciplinary rounds, and family conferences, nurses ensure that all team members are informed about the patient’s condition, progress, and discharge plan. To date, the adoption and integration of evidence-based nurse-led TC models has not been widely recognized within the Tanzania’s healthcare system. There is lack of focus on TC and few healthcare professionals are trained in chronic illness care (35). Similarly, the healthcare system lacks structured and well-defined TC programs specifically designed to address the needs of patients with chronic illnesses. To address these challenges, there is an urgent need to establish structured TC programs which addresses the healthcare needs of patients and their caregivers during hospital-to-home transitions.

### 3.1 Study limitation and mitigation

This study is prone to temporal changes, such as improvements or changes in care processes over time that might confound the observed before-and-after differences. Also, extreme values in the “before” measurement may regress toward the mean in the “after” measurement purely by chance, creating the illusion of improvement due to the intervention. However, we will use various mitigation strategies to address the limitations of this study. First, we will include multiple follow-up points to minimize the impact of short-term fluctuations that will help to differentiate sustained effects from transient effects of the intervention. Second, we will employ statistical techniques such as regression analysis to adjust for confounding variables and reduce biases. Third, we will maintain consistency in measurement tools, perform sensitivity and post-hoc analysis, and implement blinding procedures to reduce biases in outcome assessment, ensuring the outcome assessors are unaware of the participants’ intervention status.

## 4.0 Conclusion

There is a need to critically consider TC as an absolute care need for stroke survivors and their caregivers. However, the current TC in Tanzania faces several challenges such as insufficient staff training in chronic care, limited access to post-stroke care services and low patients’ health literacy. Similarly, there is a lack of structured and well-coordinated TC program that addresses the needs of stroke survivors. Thus, there’s a need for further investments in structured and locally tailored stroke TC programs, and targeted interventions to address the unique needs of stroke survivors as they move from the hospital to their homes.

## Data Availability

No datasets were generated or analysed during the current study. All relevant data from this study will be made available upon study completion.

## Acknowledgment

None

## Reporting guideline

TIDieR (Template for Intervention Description and Replication) checklist.

